# GWAS on Birth Year Infant Mortality Rates Provides New Evidence of Recent Natural Selection

**DOI:** 10.1101/2021.06.08.21258559

**Authors:** Yuchang Wu, Shiro Furuya, Zihang Wang, Jenna E. Nobles, Jason M. Fletcher, Qiongshi Lu

## Abstract

Following more than a century of phenotypic measurement of natural selection processes, much recent work explores relationships between molecular genetic measurements and realized fitness in the next generation. We take a novel approach to the study of contemporary selective pressure by examining which genetic variants are “sustained” in populations as mortality exposure declines. Specifically, we deploy a so-called “regional GWAS” that links the infant mortality rate (IMR) by place and year in the UK with common genetic variants among cohorts in the UK Biobank. These cohorts (born 1936-1970) saw a decline in IMR from above 65 per 1,000 to under 20 per 1,000, with substantial subnational variation and spikes alongside wartime exposures. Our results show several genome-wide significant loci, including *LCT* and *TLR10/1/6*, related to area-level cohort IMR exposure during gestation and infancy. Genetic correlations are found across multiple domains, including fertility, cognition, health behaviors, and health outcomes, suggesting an important role for cohort selection in modern populations.

## Introduction

A large literature over the past century has explored associations between phenotypic measures of health and social status with markers of fertility and reproductive success to draw inference about natural selection processes in the modern era^1^. These studies highlight the still-evolving nature of human populations in low-mortality settings; they also facilitate prediction about short-run population change^2^. Recent work provides new evidence on selection by leveraging large, biobank-scale genetic information, alongside new methods to summarize genome-wide measurements into polygenic scores^3^ and tests of associations between summarized genetic information and reproductive success^4,5^. For example, studies have demonstrated associations between polygenic scores for specific phenotypes and reproductive success in the U.S. and in Iceland^6-8^. Others have demonstrated single nucleotide polymorphism (SNP) correlations with fertility markers through genome-wide association studies (GWAS)^9^. Still others have established genetic correlations between a broad set of phenotypes and fertility^10^. For example, Sanjak et al. (2018)^10^ finds genetic correlations between reproductive success and age at first birth, age at menarche, age at menopause, educational attainment (EA) and cognition, as well as body mass index (BMI) and related metabolic measures. In general, the findings are of modest directional selection that would accumulate into noticeable effects on larger time scales^8^.

We advance this field by taking a novel, complementary approach to the study of selective pressure. We ask which SNP alleles survive in populations when cohorts are exposed to better disease environments—or in contrast, which alleles “disappear” in particularly harsh environments. We do this by using a “regional” GWAS^11^. We measure the disease environment experienced by cohorts during gestation and in infancy, two periods in the life course prior to reproduction when cohort mortality is comparatively high^12,13^. We index the disease environment with the prevailing infant mortality rate (IMR) by place and year. The IMR is a well-established indicator of area-level conditions related to nutrition, infection, and inflammation, particularly during the early-to mid-twentieth century^14-16^.

The approach builds on decades of demographic and epidemiologic research that considers how cohort traits are shaped by mortality exposure early in life^17-21^. These studies provide several types of indirect evidence of early-life cohort selection: the differential survival of robust subpopulations through inhospitable disease environments during gestation and infancy (e.g., female versus male survival^22,23^); nonlinear associations between disease environments and phenotypic traits like birth weight or height^24^; and phenotypic traits among *descendants* of people exposed to war and famine in early life^25,26^. We shed new light on these processes by explicitly examining molecular genetic information (SNPs) among cohorts who survive similar periods of hardship—and by testing how surviving SNPs are correlated with an array of complex traits, including the phenotypes explored in these previous studies.

We begin by examining associations between the early-life disease environment and the presence of specific loci among surviving cohort members. We then consider year-specific variation, including that aligned with particular hardships occurring during World War II. We conduct several sensitivity tests to rule out alternative explanations for the patterns detected in the UK Biobank (UKB) sample. We then examine the association between detected SNPs and the genetic predictors of an array of complex traits. We demonstrate that the shifts in genetic selection associated with improving early-life conditions are associated with genetic predictors of an array of reproductive, health, and behavioral traits.

## Results

### Infant mortality data

Area level infant mortality fell over 60% over the time period of the UKB respondents’ birth years, between 1936-1970. **Figure 1** shows the time trend and also the across-county variation in each year (also see **Supplementary Figure 1**). These reductions were the result of improved living conditions and widespread public health efforts, among them, the expansion of prenatal and infant health care^27^ after the war. Significant regional variation persisted through this period^28,29^. The IMR is highest in 1940 and 1941 when the UK population was exposed to intensive bombing campaigns during the World War II “Blitz”, alongside ongoing wartime reductions in nutrition that were particularly acute for unemployed and poor households^30,31^.

**Figure 1.**
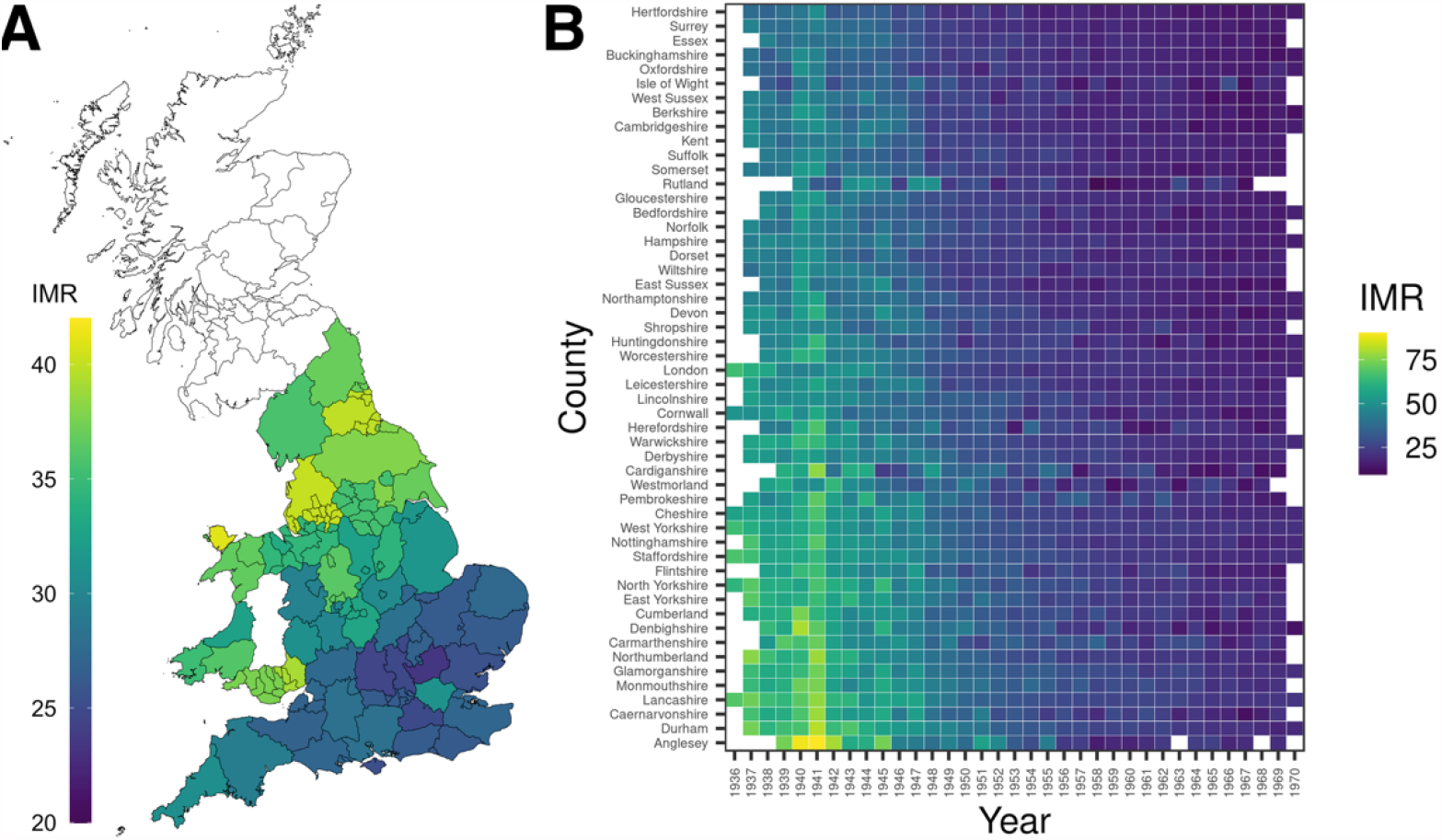
IMR between 1936-1970 in England and Wales. **(A)** Heatmap of average county-level IMR between 1936-1970 in England and Wales. IMR is defined as the probability of deaths under one year of age (per 1,000 livebirths). The denominator of the IMR does not include stillbirths. **(B)** Heatmap of IMR in each county and year.

We measure IMR to capture the disease environment in the year prior to birth and during the year of birth in the county in which each participant in the UK Biobank was born. We separately test effects of these two exposures. The “lagged” IMR—or IMR during gestation—indexes conditions related to pregnancy survival in cohorts. The birth year IMR indexes conditions related to infant survival in cohorts. Throughout the study we show findings from analysis of the birth year IMR and refer to the findings of the analysis of gestation-year IMR, which are presented in the **Supplementary Material**.

### GWAS identifies genetic loci associated with birth year IMR

We conducted a GWAS on birth year IMR using 291,591 independent UKB respondents of European descent. To adjust for population stratification and the non-linear time trend in IMR, we performed GWAS using BOLT-LMM^32^ with year-of-birth effects included as indicator variables along with other covariates (**Methods**). Following previous work^11^, we adjusted standard error of SNP effects using the intercept from linkage disequilibrium (LD) score regression^33^ to conservatively control type-I error.

We identified two loci reaching genome-wide significance (**Figure 2** and **Table 1**): the lactase (*LCT*) locus on chromosome 2 (rs1446585; p = 1.3e-17) and the *TLR1*-*TLR6*-*TLR10* gene cluster on chromosome 4 (rs5743618; p = 1.8e-13). Another locus near the *EFTUD1* gene (a.k.a. *EFL1*) on chromosome 15 showed a suggestive association (rs9944197; p = 6.6e-8; **Supplementary Figure 2**). SNP heritability was low but statistically significant (h^2^ = 0.016, SE = 0.002), with an inflation factor *λ* = 1.06 (**Supplementary Figure 3**). Heritability showed a two-fold depletion in repressed genome but did not reach statistical significance after correcting for multiple testing (**Supplementary Table 1**).

**Figure 2.**
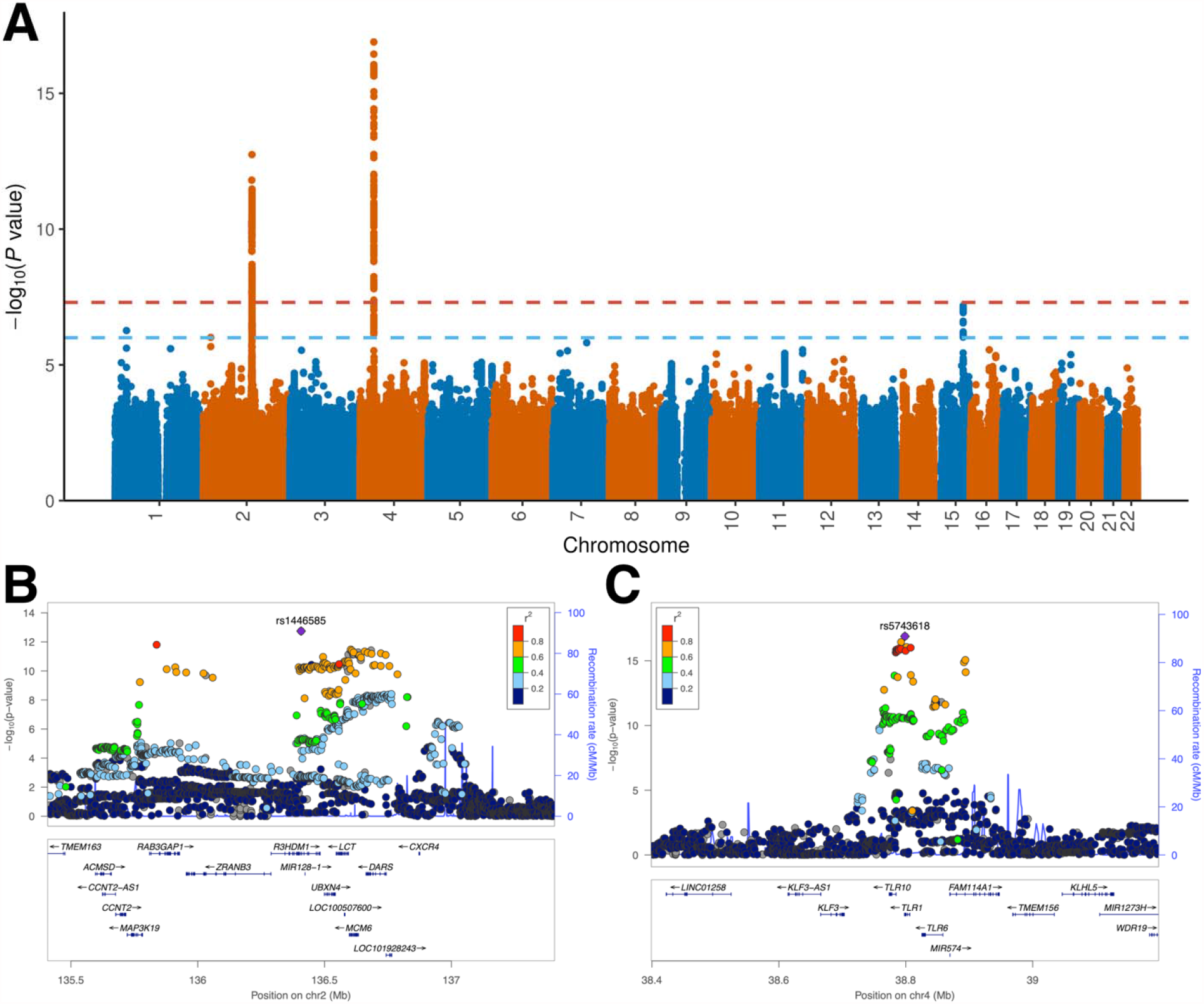
Genetic associations for birth year IMR. **(A)** Manhattan plot for birth year IMR. The horizontal lines mark the genome-wide significance cutoff of 5.0e-8 and a suggestive cutoff of 1.0e-6. **(B)** Genetic associations at the *LCT* locus. **(C)** genetic associations at the *TLR1/6/10* locus.

**Table 1.**
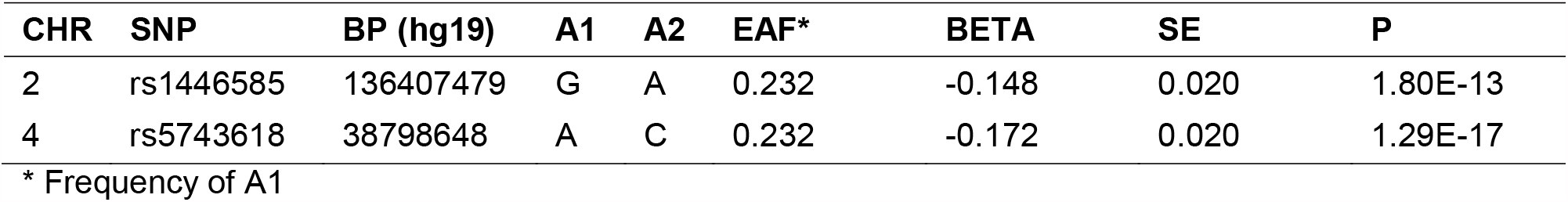
Genome-wide significant loci associated with birth year IMR.

Conceptually, in order to focus on measures of the disease environment in the *in utero* period of each respondent, we use the IMR in the year prior to birth to capture the prevailing nutrition and infection conditions. We refer to this as the lagged-IMR phenotype. In practice, using the lagged or contemporaneous measure of IMR produces results with a genetic correlation of 1.002 (SE = 0.016; **Supplementary Figures 4-5**).

Our findings are unlikely due to participation bias in the UKB^34,35^. Adjusting for participation activities in GWAS and conditioning on a latent proxy factor for UKB participation (**Methods**) both yielded highly consistent association results (**Supplementary Figures 6-8**). In order to further guard against spurious findings, we also conducted falsification tests, where we randomly shuffled the place of the respondent’s IMR measurement. We found null results (**Supplementary Figures 8-9**).

### GWAS findings suggest recent selection in the UK

Our GWAS on birth year IMR identified two associated loci, i.e. *LCT* and *TLR1/6/10*, both of which are well-known targets of selection in Europeans^36-41^. The beneficial alleles at both loci are associated with higher birth year IMR in our analyses, which is consistent with positive selection of these alleles in tougher environments. A recent study of more than 200 ancient genomes identified 12 target loci, including both *LCT* and *TLR1/6/10*, with strong signals of selection possibly through their associations with nutrition, immunity, pigmentation, and other human traits^37^. We assessed associations between these loci and birth year IMR (**Figure 3a**; **Supplementary Table 2**). Genetic loci for pigmentation showed substantially weaker associations with IMR compared to loci associated with lactase persistence (*LCT*), resistance to leprosy and other mycobacteria (*TLR1/6/10*), and vitamin D metabolism (*DHCR7* and *NADSYN1*). Additionally, the strength of IMR GWAS associations are correlated with IMR-increasing single density scores (SDS; Spearman correlation = 0.036, p = 9.6e-13; **Figure 3b**; **Methods**), suggesting polygenic and positive selection on IMR-associated alleles. We also confirmed this relationship using bivariate LD score regression (**Supplementary Figure 10**; genetic correlation = 0.25; p = 0.01). Notably, the *LCT* locus strongly colocalizes with high SDS while the signal at *TLR1/6/10* was not as strong (**Supplementary Figure 11**).

**Figure 3.**
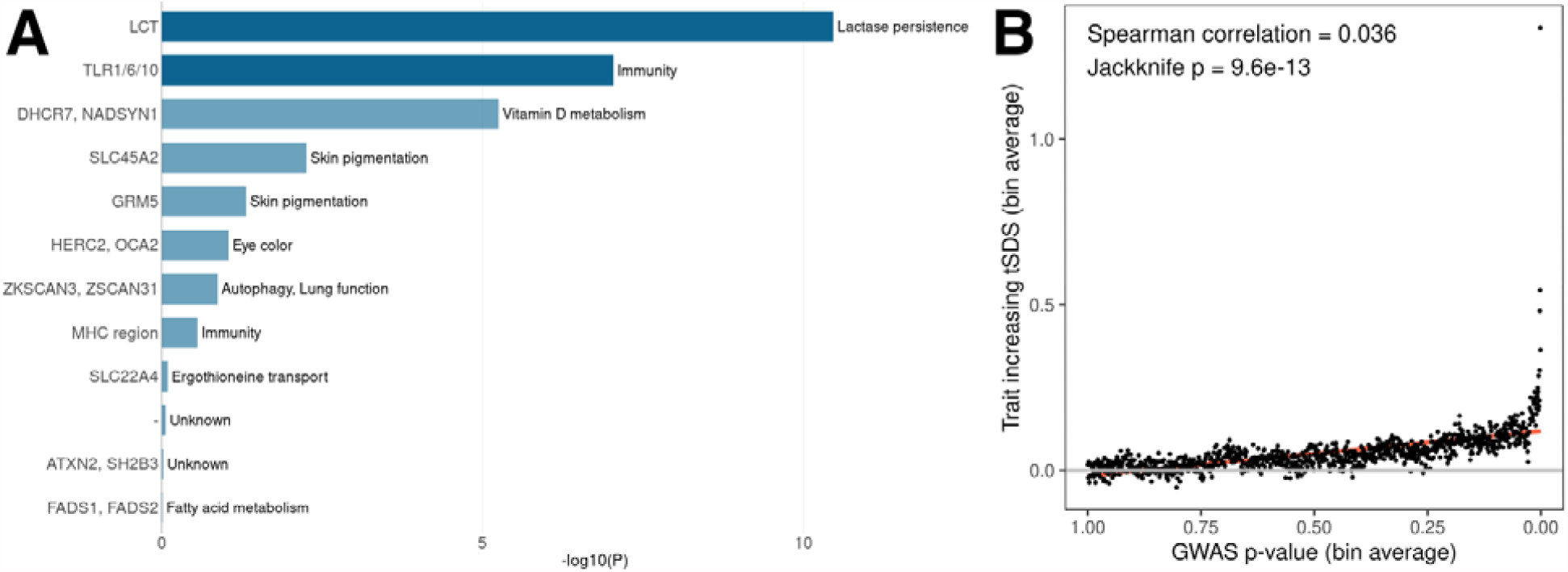
Selection patterns at IMR-associated genomic loci. **(A)** Associations with birth year IMR at 12 target loci for selection in Europeans (also see **Supplementary Table 2**). Genetic loci reaching genome-wide significance in the IMR GWAS are highlighted in dark blue. **(B)** Correlation between GWAS associations of birth year IMR and SDS matched with IMR-associated alleles.

We further partitioned UKB samples by year of birth and estimated SNP-IMR associations for each year separately. Effect sizes at both the *LCT* and *TLR1/6/10* loci peak at 1941, during “The Blitz” (**Figure 4a**). In particular, the *LCT* locus (rs1446585) showed substantially attenuated associations with birth year IMR before 1940 and after 1942. We compared the allele frequency of rs1446585 in samples born in the top 10 counties with the highest IMR and in the 10 counties with the lowest IMR. We found substantial differences in allele frequencies between counties with high and low IMR. In counties with high IMR, the lactase-persistence-associated allele of rs1446585 (major allele) showed significantly increased frequency in the 1940 birth cohort (p = 0.043), maintained similar frequencies through 1940-1942, and had significantly reduced frequency in samples born in 1943 (p = 0.032; **Figure 4b**). The year-to-year comparison did not reach statistical significance in counties with low IMR (**Supplementary Table 3**). We found null results for the *TLR1/6/10* locus (**Supplementary Figure 12**). We also note that the substantially elevated IMR in 1940 and 1941 is not correlated with the density and frequency of bombing events (**Supplementary Figure 13**) and may be instead explained by other factors such as food scarcity during the war.

**Figure 4.**
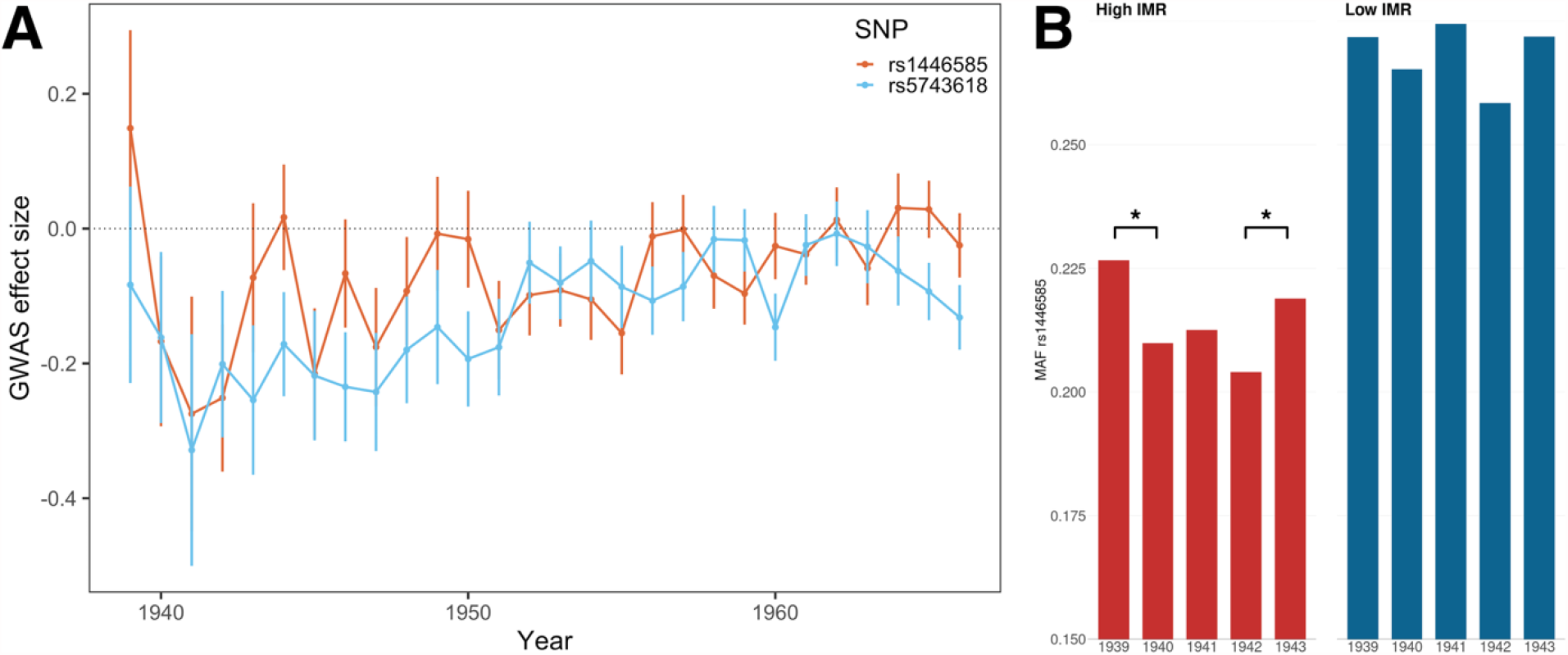
Selection on *LCT* and *TLR1/6/10* loci during “The Blitz”. **(A)** The effect size of lead SNPs at both genome-wide significant loci peak in 1941. Years with N<5000 were excluded from the analysis. **(B)** Minor allele frequency of rs1446585 in UKB birth cohort in 1939-1943. Major allele at this locus is known to associate with lactase persistence. Counties with high/low IMR were defined based on IMR in 1939.

### Genetic correlation with 50 complex traits

We next examine genetic correlation between birth year IMR and a set of 50 traits widely assessed as outcomes of selection processes (**Figure 5**; **Supplementary Table 4**). Among known target traits of selection^37^, we found a significant correlation with vitamin D but found null results on hair and skin color. Our results are consistent with other approaches showing correlations with genetics of fertility (number of children, age at first birth) but do not find effects for age at menarche or age at menopause. Recall that Sanjak et al. (2018) reported inconsistent findings between reproductive success and age at menarche (positive) and age at menopause (negative), which the authors label as “less explicable” than other results^10^. Similar to earlier findings^6,8^, we show correlations with EA and cognition, but extend this finding by showing these results are driven by direct-EA component and not by indirect-EA component (i.e., genetic nurture) using methods in Wu et al. (2020)^42^. The difference in these findings suggest a broader need for caution when examining the genetic correlation findings, as we cannot decouple parental and child genetics in these results. We also find relationships with anthropometrics, like Sanjak et al. but substantially extend our domains of interest to show novel findings for cardiovascular disease, tobacco use, and a variety of mental health conditions. The null findings on birth weight are suggestive that studies linking birthweight to insults akin to those prevailing during the 1930s and 1940s in the UK are likely capturing the deleterious effects of the disease environment (and accompanying wartime conditions) during that time, versus the differential survival of pregnancies^43,44^.

**Figure 5.**
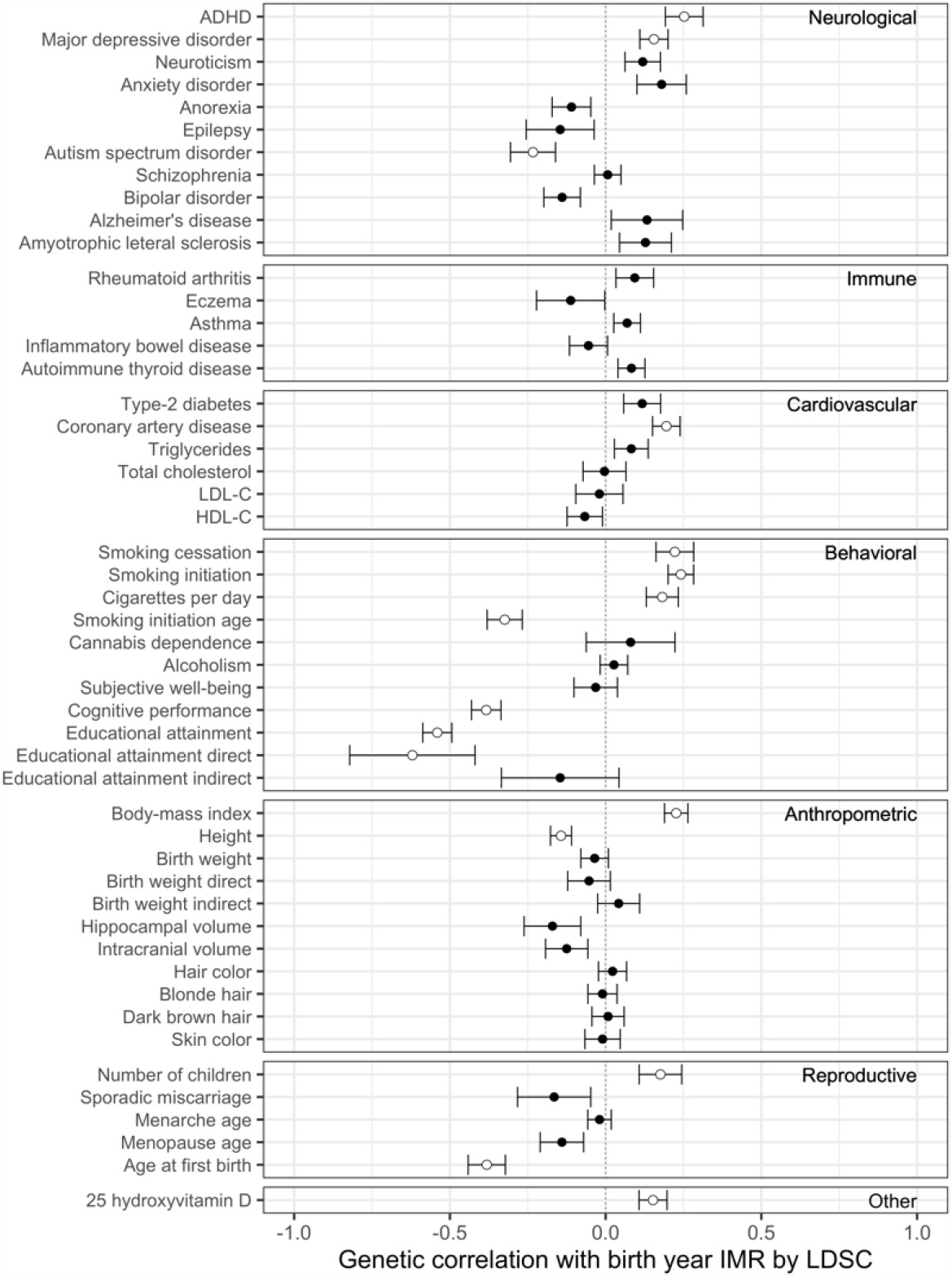
Genetic correlation between birth year IMR and 50 complex traits. Dots and intervals indicate genetic correlation estimates and standard errors. Significant correlations at a false discover rate (FDR) cutoff of 0.05 are highlighted with circles. ADHD: attention-deficit/hyperactivity disorder; LDL-C and HDL-C: low (high) density lipoprotein cholesterol.

## Discussion

In this work, we implemented a novel approach to studying natural selection in contemporary populations. We used GWAS to estimate how the frequency of common SNPs vary with area-level measures of infant mortality during the *in utero* period of UKB respondents and found two genome-wide significant signals at *LCT* and *TLR1/6/10*. These loci accord with previous work on natural selection comparing ancient and modern populations^37^. We found limited evidence of large effects across the genome and estimated SNP heritability to be less than 2%. We then show moderate genetic correlations between the IMR-GWAS and a host of phenotypic domains, reinforcing earlier findings related to fertility (number of children, age at first birth), anthropometrics (BMI, height), and cognition (EA, fluid intelligence) and also extending findings into psychiatric conditions (major depressive disorder, ADHD, autism) and health conditions (coronary artery disease).

Compared to past studies comparing allele frequencies in ancestral and modern populations^37,45^ and inferring lengths of the genealogy^38,46^, this study directly estimates the shift of allele frequencies in less favorable environments. It identifies specific genomic loci under very recent selection and provides fundamental new insights into the mechanism and timing of such selection. We found evidence for selection on lactase and potential resistance to leprosy and other mycobacteria in the past century and found null results for pigmentation traits. In particular, we demonstrate accelerated selection on these specific alleles in the UK during the mortality conditions caused by the World War II bombing campaigns during the Blitz.

We also note that because our analysis is tied to area-level infant mortality, it differs from analysis that links reproductive success (number of children born) with genetic measurement^9,10^. In part, this is because the parents of the cohorts studied here who lose a pregnancy or an infant during periods of high infant mortality may have a subsequent successful pregnancy and surviving infant and thereby achieve levels of reproductive success. Instead, the focus here is on the genetic characteristics of people born between 1936-1970 who survive through reproductive age. We find that these survivors have genetic traits that are correlated with early initiation of childbearing and a larger number of children. Like past research, we generally are unable to separate child (direct) and parental (indirect) genetic relationships^47^ in the analysis. The results for EA suggest the effects are direct, which suggests that selection is occurring at the level of the child’s genetics rather than through family-level correlates of child bearing and survival, such as socioeconomic status.

Our study has a few limitations. We conducted analyses in UKB and it is unclear if these findings can be replicated in other populations. Second, although we provide evidence on the timing of selection and its ongoing nature, the specific mechanisms underlying such selection remains unclear. Even in the World War II example, we cannot parse out effects of stress, nutrition, or other related mechanisms. Although we demonstrate that these findings are unlikely due to participation bias, we cannot distinguish infant death or impacts on health later in life. Further, we deployed state-of-the-art approaches (e.g., BOLT-LMM and LD score regression-based genomic control) to adjust for population stratification in GWAS, but applications involving polygenic modeling (e.g., polygenic scores) may still be susceptible when weak but systematic bias is aggregated across many SNPs. Recent results suggest that UKB is relatively less susceptible to population stratification compared to other cohorts^48,49^ and methods for heritability and genetic correlation estimation (e.g., LD score regression) are robust to unadjusted population stratification in GWAS^50^. Still, associations in IMR GWAS need to be interpreted with caution. Another limitation facing most current research using genetic information is the focus on respondents of European ancestry. This study also faces this key limitation. Approximately 10% of the UKB sample was born in the UK but has African, Asian, or other ancestry. Data to support estimation of GWAS in populations with ancestry outside of Europe is growing and much needed. At present, the findings of this study are limited in applicability to the UK population with European ancestry.

Taken together, the research makes multiple contributions to the study of selective pressure using molecular genetic data. For example, several landmark studies^37,38,46^ have found specific genomic loci under selection over the time span of several millennia. The approach we use here, a regional GWAS, is designed to be exploratory and hypothesis-free, facilitating detection of previously undescribed relationships for contemporary cohorts. It is remarkable then, that the approach in this study finds evidence of selection on the same loci as previous explorations using ancient genomes. In doing so, we demonstrate that selection described over the long arc of human history is occurring in the contemporary era. Further, we aggregated SNPs into domains of phenotypes through genetic correlation assessments and demonstrated that the shifts in genetic selection associated with improving early-life conditions are associated with an array of reproductive, health, and behavioral traits. These results shed important light on how selective pressure is correlated with complex traits in contemporary populations.

## Methods

### IMR data in England and Wales

Regional IMR were obtained from A Vision of Britain through Time (https://www.visionofbritain.org.uk), which includes vital statistics for annual counts of infant deaths and total births for each of over 400 local government districts in England and Wales. We created a crosswalk between the local government districts and the counties in the 2011 Census in order to merge the IMR to the place of birth information in the UKB.

For visualization purpose, we downloaded maps and spatial data for the UK from “GADM” website (https://gadm.org/download_country_v3.html). We used the “sf” R package to process spatial data and plotted maps.

### GWAS analysis in the UKB

Following Abdellaoui et al^11^, we deployed a regional GWAS in UKB to test associations between genetic measures at the individual level and IMR, where all subjects reporting the same place of birth (data fields 129/130) and same year of birth (data field 33) had the same regional phenotypic value assigned. Of the 500,000 participants in UKB, we focused on the respondents with European ancestry and those with places of birth in England and Wales (data field 1647) in order to match with our contextual data. We excluded the participants who are recommended by UKB to be excluded from analysis (data field 22010), those with conflicting genetically-inferred (data field 22001) and self-reported sex (data field 31), and those who withdrew from UKB. UKB samples with European ancestry were identified from principal component analysis (data field 22006). We used KING^51^ to infer the pairwise family kinship among UKB samples and identified 154 pairs of monozygotic twins, 242 pairs of fraternal twins, 19,136 full sibling pairs, and 5,336 parent offspring pairs among 408,921 individuals of European descent. A total of 291,591 independent samples born between 1936 and 1970 were used in the IMR GWAS. We used BOLT-LMM^32^ to perform GWAS with sex, genotype array (data field 22000), and year of birth as covariates. Year of birth was dummy coded to account for the nonlinear time trend in IMR (**Supplementary Figure 1**). We kept only the SNPs with missing call rate ≤ 0.01, minor allele frequency 2: 0.01, and with Hardy Weinberg equilibrium test p-value ≥ 1.0e-6 in GWAS. We also applied LD score regression-based genomic control^11,33^ where we inflated the standard error of SNP effects using 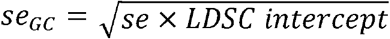 to conservatively control unadjusted confounding in association tests.

### Heritability and genetic correlation estimation

We used LD score regression^52^ implemented in the LDSC software to estimate heritability of IMR, quantify heritability enrichment in 52 baseline functional annotations, and estimate genetic correlations of birth year IMR with 50 complex traits using GWAS summary statistics as input. Details of the 50 traits used in genetic correlation analysis are shown in **Supplementary Table 4**.

### Quantifying and removing participation bias

We performed two secondary analyses to account for participation bias in GWAS. First, we conditioned the analysis on participation status of the optional mental health questionnaire (MHQ) in UKB. We categorized GWAS samples into 3 strata: MHQ = 0 (received the invitation but did not participate; N = 97,401), 1 (received the invitation and participated; N = 95,667), or “NA” (did not receive the invitation due to unavailable email addresses; N = 98,523). Samples who did not receive invitations are enriched for older age lower education^53^. Within each MHQ stratum, we performed GWAS on birth year IMR using the same settings described above for the main GWAS.

We also used genomic structural equation modelling (GSEM)^54^ to remove participation bias. Our model jointly regressed three GWAS of participation of optional questionnaires, i.e. MHQ, food frequency questionnaire (FFQ), and physical activity study (PAS) in UKB on a latent factor F representing participation behavior (**Supplementary Figure 7**). The latent factor F and IMR were then regressed on each SNP to estimate participation-adjusted genetic association with birth year IMR. Since the coefficients and standard errors in GSEM were on the scale of standardized phenotypes, we transformed GSEM results to the same scale of BOLT-LMM outcomes for comparison.

### SDS data and analysis

SDS were computed using 3,195 individuals from the UK10K project^38^. To match alleles between SDS and IMR GWAS, we first chose effect alleles in GWAS to always have positive associations with birth year IMR. Then, we obtained trait-SDS (tSDS) by transforming SDS such that tSDS = SDS if the derived allele in SDS is the effect allele in GWAS and tSDS = -SDS if the derived allele in SDS is the non-effect allele in GWAS. We set tSDS to be “NA” if otherwise. Following Berg et al.^48^, we estimated Spearman correlations between SDS and GWAS z-scores and used a block-jackknife approach to obtain the standard errors and p-values for Spearman correlations. To remove spurious correlations caused by unadjusted confounding (e.g., population stratification), we also applied LD score regression to compute genetic correlations between SDS and IMR GWAS associations.

### Data on air raids during World War II

Detailed data on German air raids on the UK during World War II were obtained from (http://www.warstateandsociety.com/Bombing-Britain). We accessed the coordinates of bombing events and marked their locations on the map of UK using the “ggmap” R package^55^. We defined bombing density as the number of bombing events with casualty during 1940-1942 within a 10 km radius of each UKB participant’s place of birth.

## Data Availability

Data available from UK Biobank

## Data availability

GWAS summary statistics for birth year IMR are available at (http://qlu-lab.org/data.html).

## Acknowledgements

The authors gratefully acknowledge use of the facilities of the Center for Demography of Health and Aging at the University of Wisconsin-Madison, funded by NIA Center Grant P30 AG017266. We thank members of the Social Genomics Working Group at University of Wisconsin for helpful comments. This research has been conducted using the UK Biobank Resource under Application 57284.

## Author contribution

J.F. and Q.L. conceived and designed the study.

Y.W. and Q.L. performed the analyses.

S.F. obtained and processed IMR data.

Z.W. assisted in analyzing and visualizing World War II air raid data and performed GSEM analyses.

J.N. advised on IMR literature and interpretation.

Y.W., J.N., J.F., and Q.L. wrote the manuscript.

All authors revised and approved the manuscript.

