## Supplementary Figures for "GWAS on Birth Year Infant Mortality Rates Provides New Evidence of Recent Natural Selection"

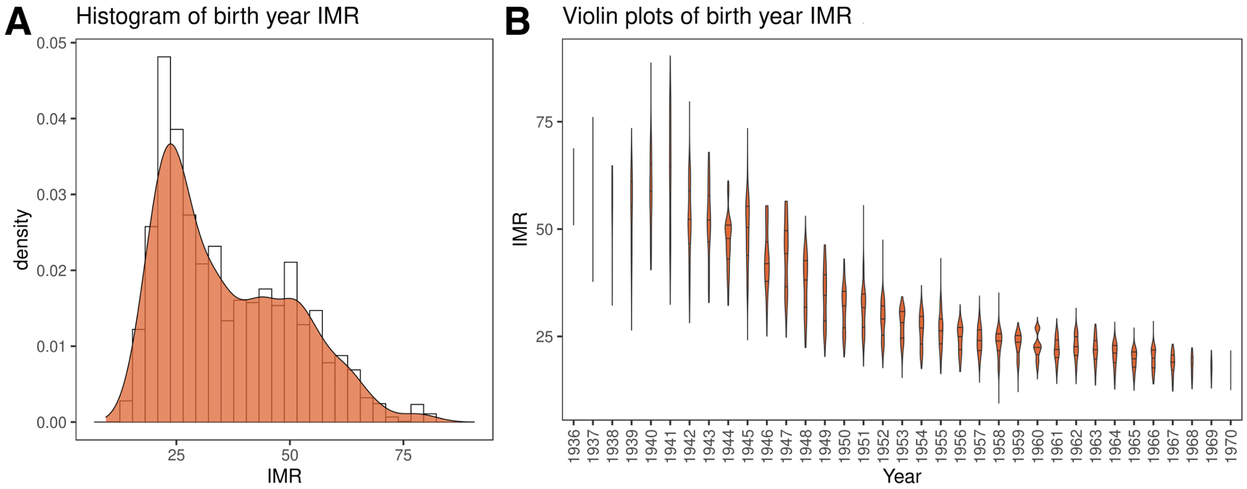


**Supplementary Figure 1. Distribution of birth year IMR across counties and year. (A)** Distribution density of birth year IMR in UK Biobank samples. **(B)** Violin plot of birth year IMR stratified by year of birth.

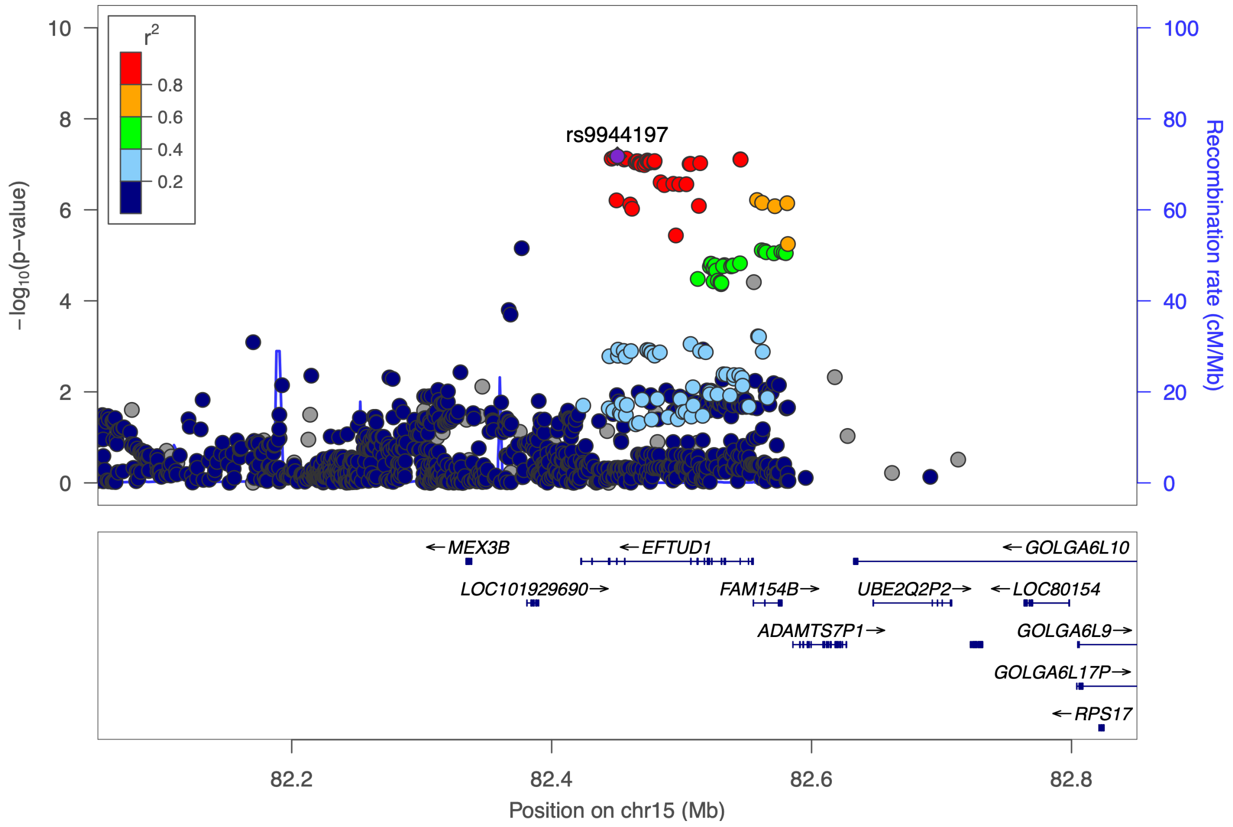


**Supplementary Figure 2. Genetic associations with birth year IMR at the *EFTUD1* locus.**

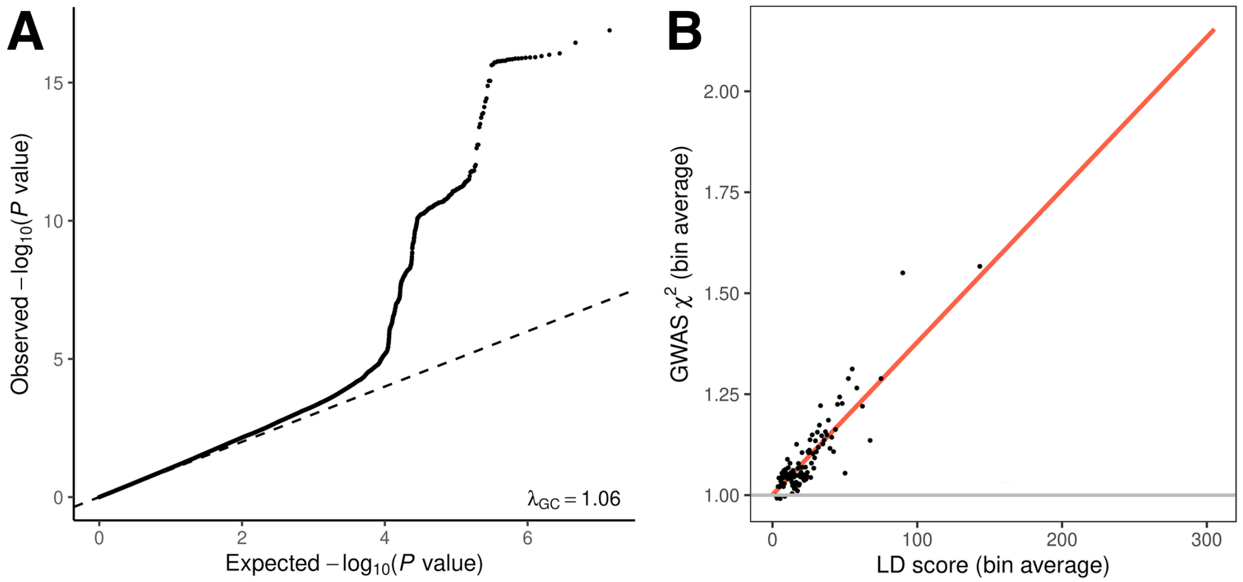


**Supplementary Figure 3. Polygenic genetic basis of birth year IMR. (A)** QQ plot for GWAS of birth year IMR. **(B)** LD score regression suggests a positive heritability of birth year IMR. We grouped SNPs into 100 bins based on LD scores. Each point in the plot shows the average LD score and the average GWAS chi-square statistics in a bin. Only HapMap 3 SNPs were included in the analysis.

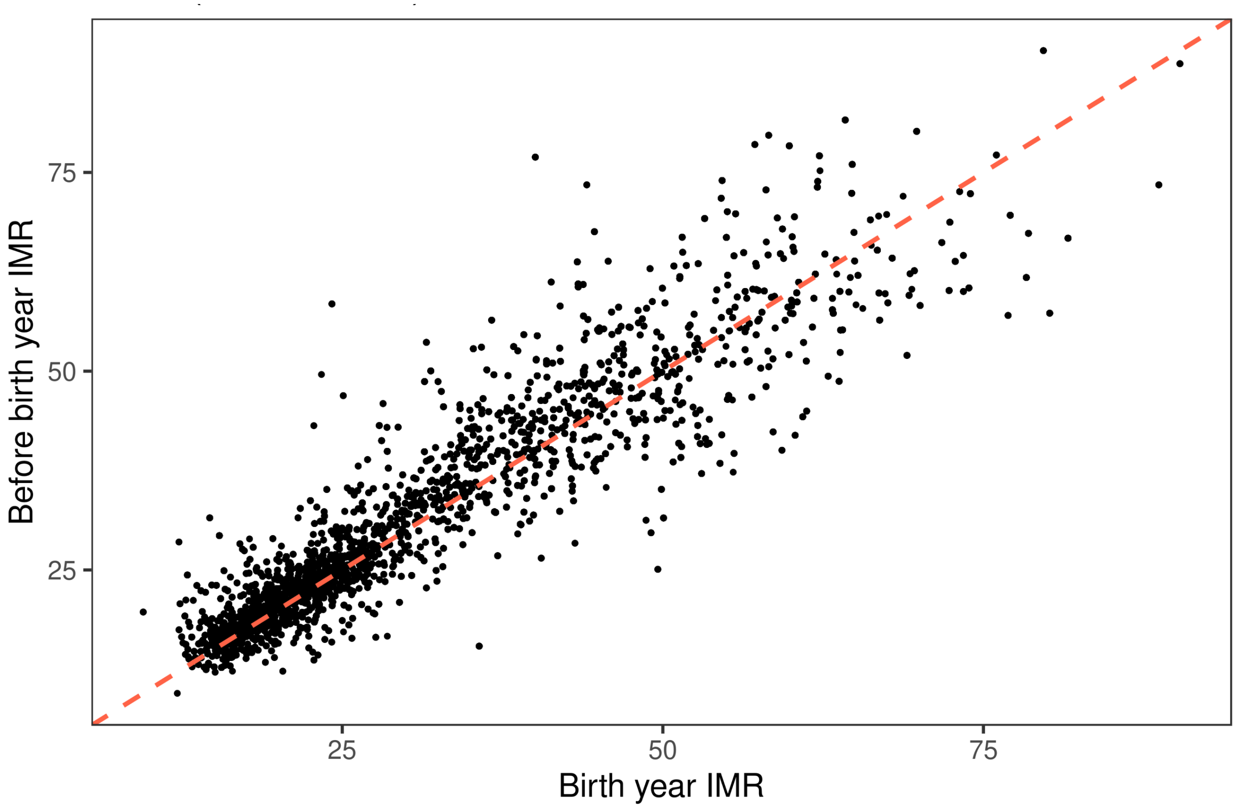


**Supplementary Figure 4. Correlation between birth year IMR and lagged-IMR in UKB samples.**

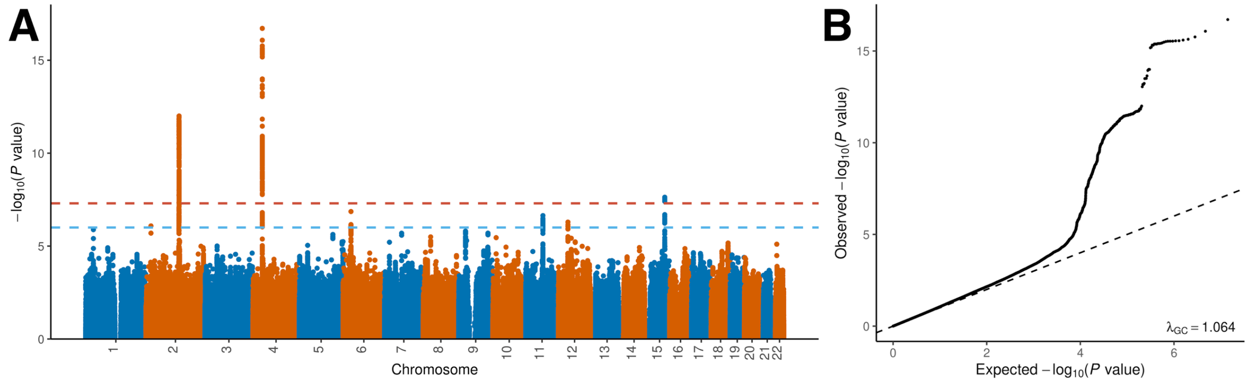


**Supplementary Figure 5. Genetic associations with IMR of the year before birth.** **(A)** Manhattan plot for lagged IMR. The horizontal lines mark the genome-wide significance cutoff of 5.0e-8 and a suggestive cutoff of 1.0e-6. **(B)** QQ plot for GWAS of lagged IMR.

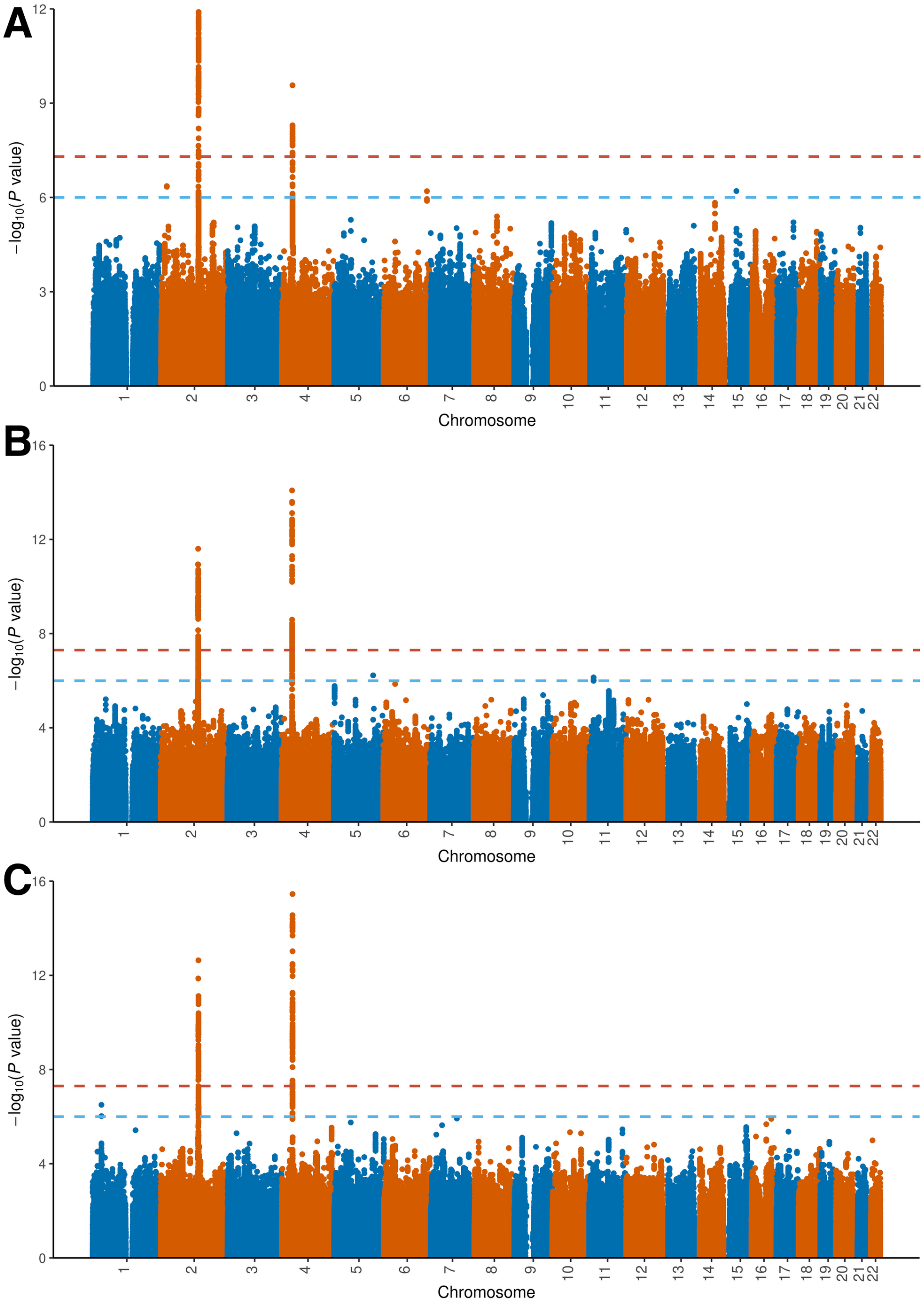


**Supplementary Figure 6. Genetic associations with birth year IMR (A) in participants of the MHQ questionnaire, (B) in non-participants of the MHQ questionnaire, and (C) after controlling for a latent factor for participation activity.** The horizontal lines mark the genome-wide significance cutoff of 5.0e-8 and a suggestive cutoff of 1.0e-6.

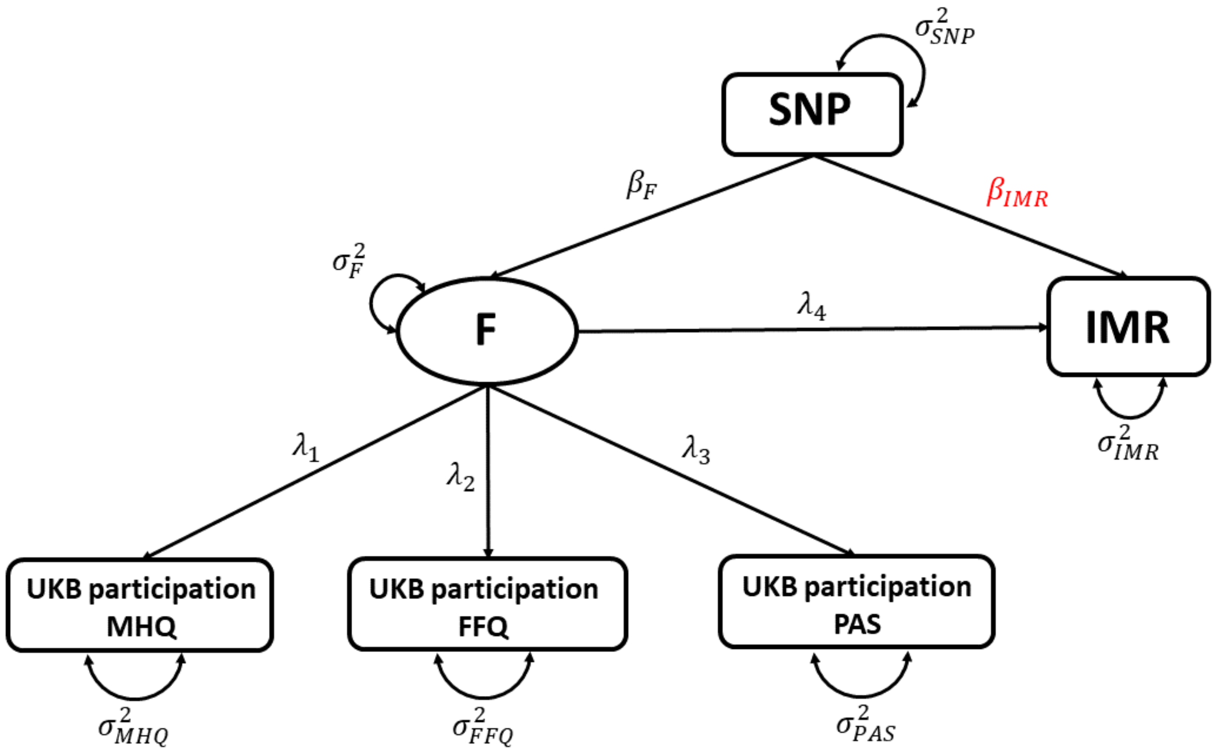


**Supplementary Figure 7. Accounting for a latent factor for participation in the GWAS of birth year IMR.** GSEM was used to fit the model for each SNP separately. The parameter of interest is highlighted in red.

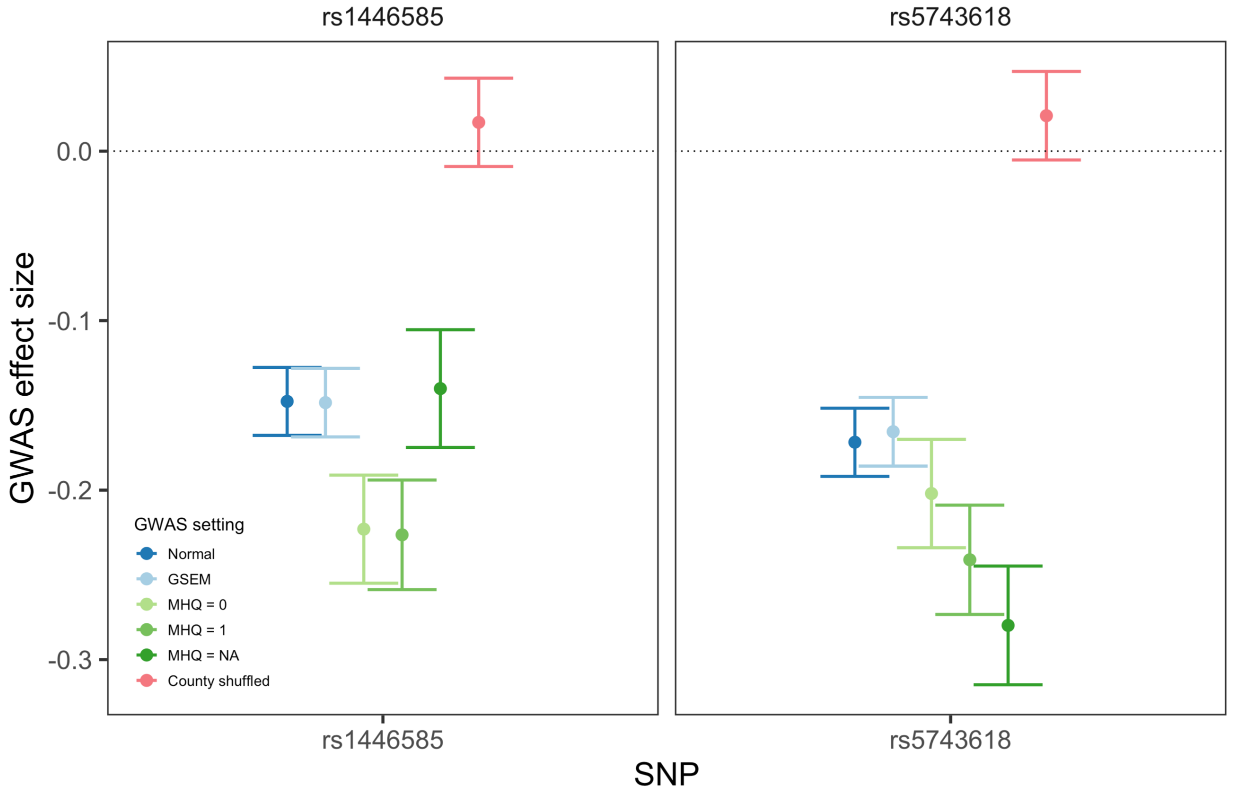


**Supplementary Figure 8. Effect size of lead SNPs at *LCT* and *TLR1/6/10* loci.** The intervals indicate point estimates and standard errors of SNP-IMR associations in the primary GWAS (dark blue), in GSEM-based analysis adjusting for participation (light blue), in analyses conditioning on participation status of the MHQ questionnaire (green), and in GWAS with county-shuffled IMR phenotype (pink).

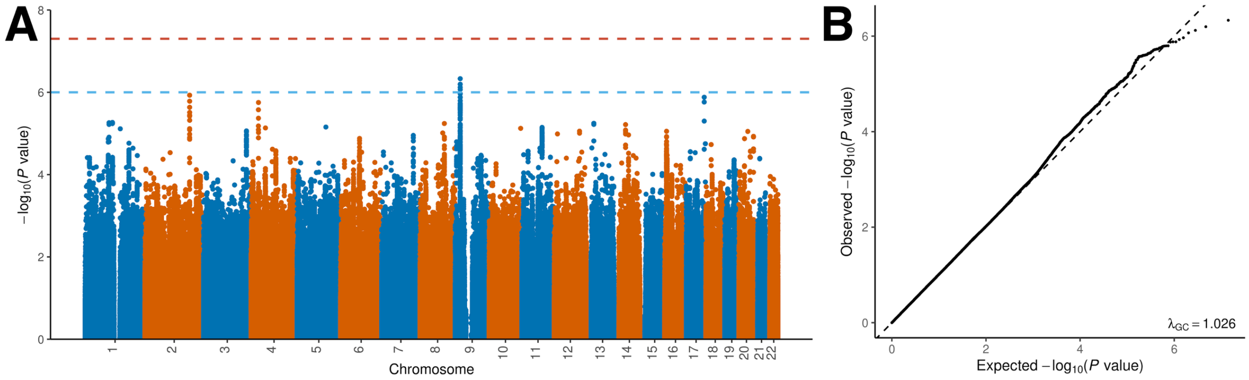


**Supplementary Figure 9. Genetic associations with birth year IMR after randomly shuffling county of birth. (A)** Manhattan plot for region-shuffled IMR. The horizontal lines mark the genome-wide significance cutoff of 5.0e-8 and a suggestive cutoff of 1.0e-6. **(B)** QQ plot for GWAS of region-shuffled IMR.

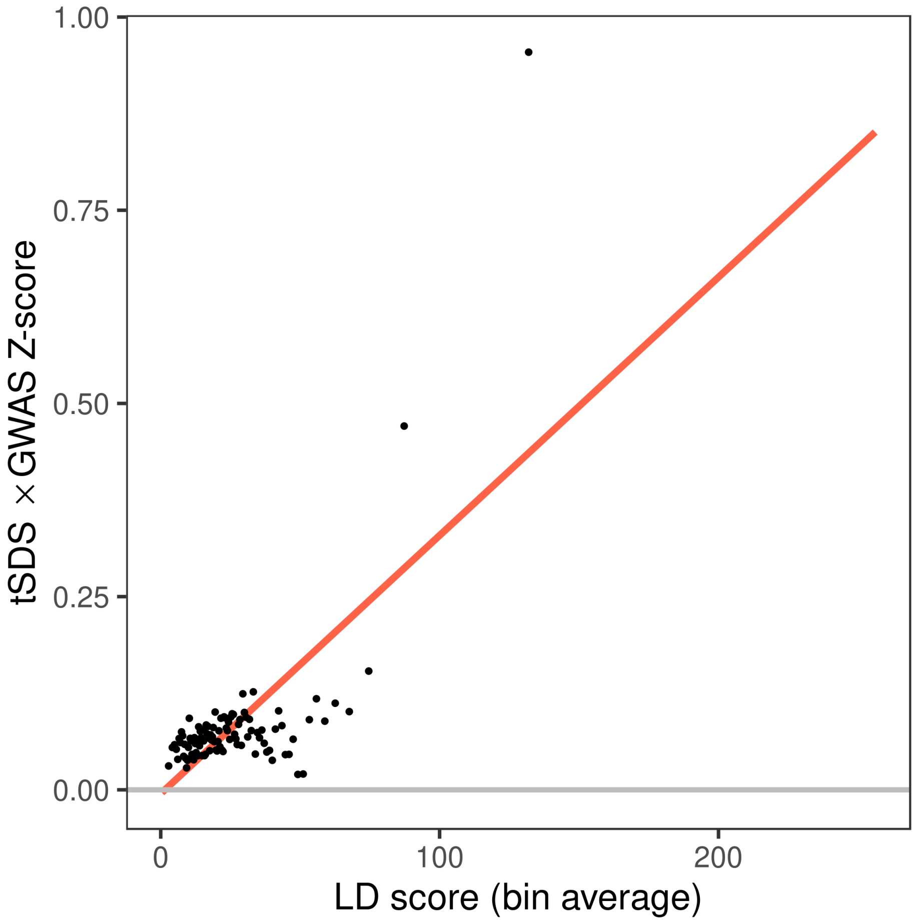


**Supplementary Figure 10. Bivariate LD score regression confirms correlation between IMR associations and SDS.** We grouped SNPs into 100 bins based on LD scores. Each point in the plot shows the average LD score and the average product of z-scores in a bin. Only HapMap 3 SNPs were included in the analysis.

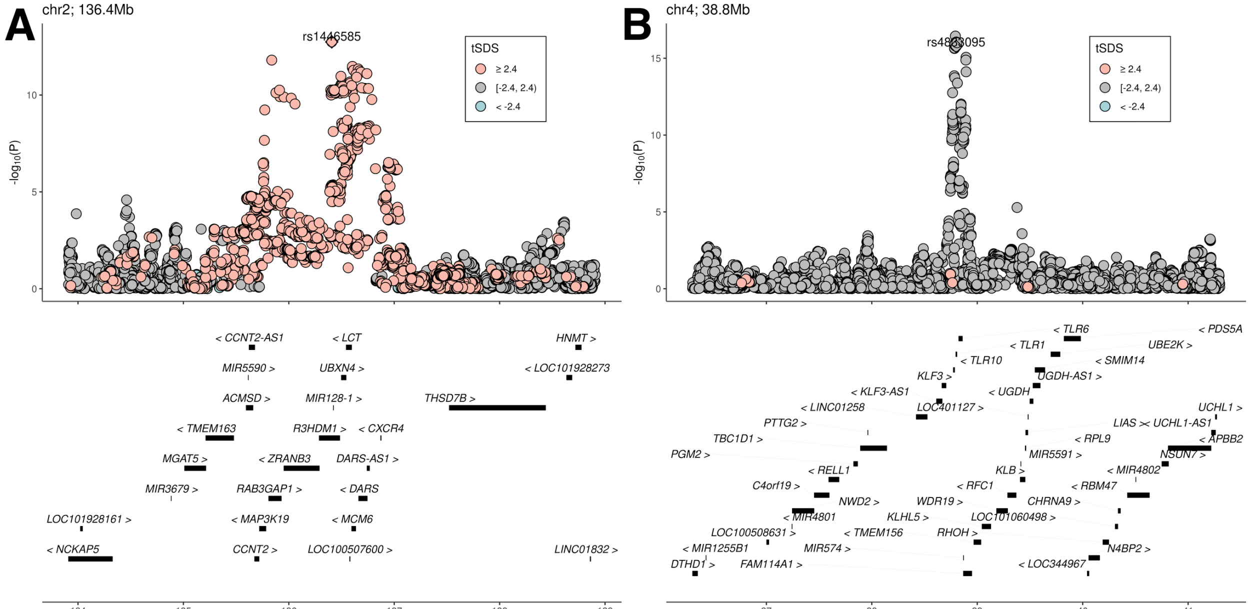


**Supplementary Figure 11. SDS values at the (A) *LCT* and (B) *TLR1/6/10* loci.** The X-axis shows hg19 genome coordinates.

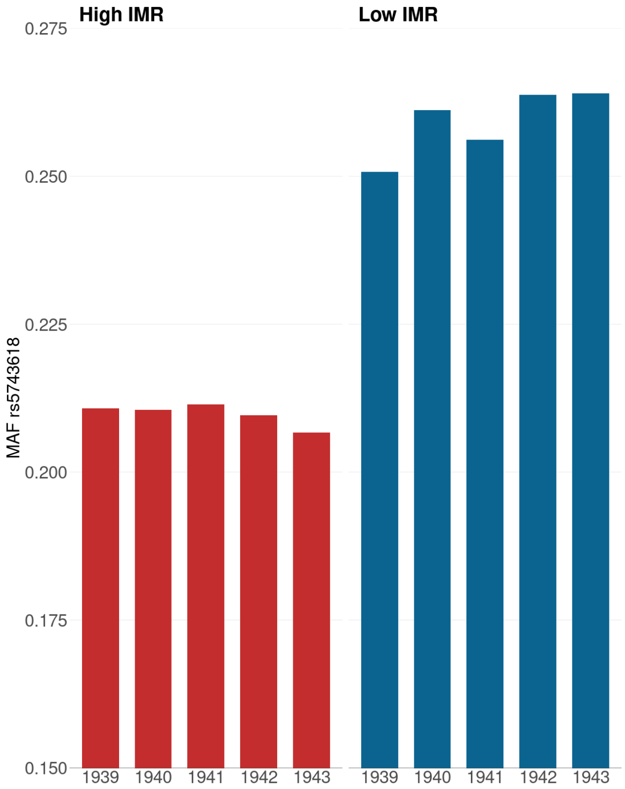


**Supplementary Figure 12. Minor allele frequency of rs5743618 in UKB birth cohort in 1939-1943.** Major allele at this locus is known to associate with resistance to leprosy.

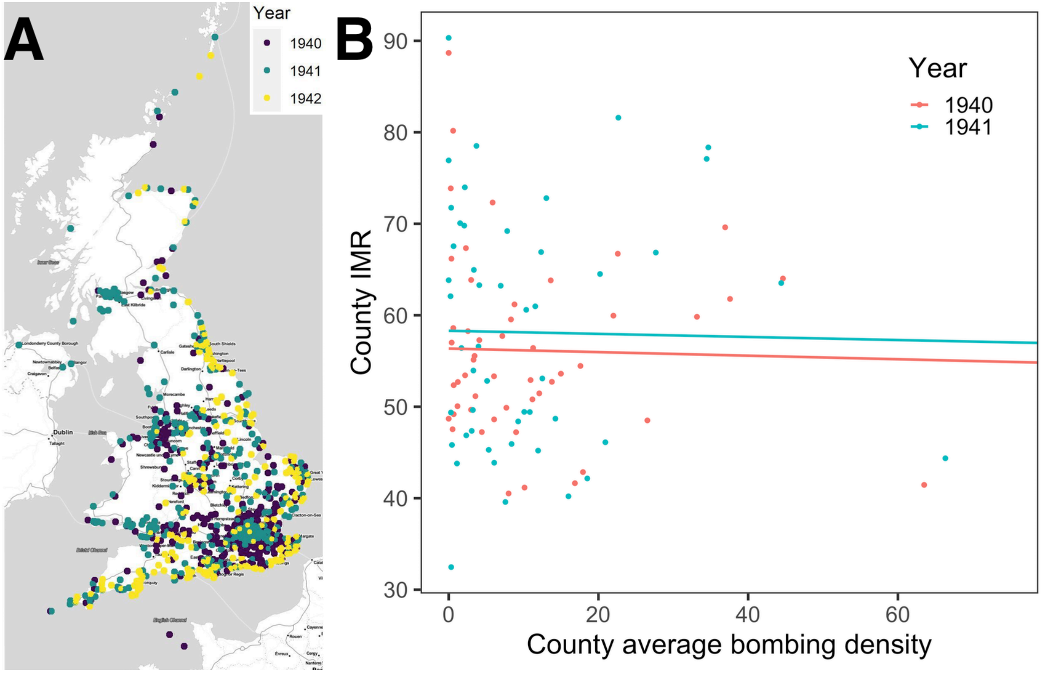


**Supplementary Figure 13. A lack of correlation between bombing density and county-level IMR during the war. (A)** Location of bombing with casualty during 1940-1942 in the UK. **(B)** Scatter plot for county-level IMR and average bombing density in 1940 and 1941. Bombing density for each UKB participant was defined as the number of bombing events with casualty in the participant's year of birth and within a 10km radius of the participant's place of birth. Values shown on the X-axis are the average bombing density for the 1940/1941 birth cohort for each county. Two counties were omitted from the plot due to very high bombing density.
